# Mammographic density assessed using deep learning in women at high risk of developing breast cancer: the effect of weight change on density

**DOI:** 10.1101/2024.06.22.24309234

**Authors:** Steven Squires, Michelle Harvie, Anthony Howell, D. Gareth Evans, Susan M. Astley

## Abstract

**Objectives:** High mammographic density (MD) and excess weight are associated with increased risk of breast cancer. Weight loss interventions could reduce risk, but classically defined percentage density measures may not reflect this due to disproportionate loss of breast fat. We investigate an artificial intelligence-based density method, reporting density changes in 46 women enrolled in a weight-loss study in a family history breast cancer clinic, using a volumetric density method as a comparison.

**Methods:** We analysed data from women who had weight recorded and mammograms taken at the start and end of the 12-month weight intervention study. MD was assessed at both time points using a deep learning model, pVAS, trained on expert estimates of percent density, and Volpara ™ density software.

**Results:** The Spearman rank correlation between reduction in weight and change in density was 0.17 (−0.13 to 0.43) for pVAS and 0.59 (0.36 to 0.75) for Volpara volumetric percent density.

**Conclusions:** pVAS percent density measurements were not significantly affected by change in weight. Percent density measured with Volpara increased as weight decreased, driven by changes in fat volume.

**Advances in knowledge:** The effect of weight change on pVAS mammographic density predictions has not previously been published.

## 1) Introduction

Overweight and obesity are associated with increased risk of breast cancer in both the general population and women with a family history of the disease (1). Weight loss programmes could thus be an important tool for reducing cancer risk. Any risk reduction because of weight loss would ideally be indicated by favourable changes in biomarkers of breast cancer risk.

Mammographic density (MD) is strongly related to risk of breast cancer, with women with higher density having a substantially increased risk (2). While MD has previously been defined as the ratio of dense (fibroglandular) tissue to fatty tissue in the breast (3) there are different techniques for measuring density, including area-based and volumetric approaches along with methods that also characterise the appearance of parenchymal tissue (4,5). Methods currently employed include subjective visual assessment into BI-RADS categories with qualitative descriptors (6) and subjective quantitative assessment of percent density recorded on Visual Analogue Scales (VAS) (7), automated estimates of volumes of fatty and dense tissue (8), and AI-based methods taking into account both quantity and pattern of dense tissue (9). The effect of weight on mammographic density may vary for these different measures of mammographic density.

Mammographic density has previously been shown to increase with reducing weight, due to reductions in fatty tissue volume that dominate any change in volume of fibroglandular tissue (10). Whilst higher breast density is associated with an increased risk of cancer, reduction in weight has been associated with a reduced cancer risk (11). Thus, there is confounding of percentage mammographic density with change in Body Mass Index (BMI) (12). Volumetric density is a better measure of risk when adjusted for BMI as well as age (13).

In this paper we investigate changes in mammographic density that occur alongside weight change using retrospective data from a weight loss study. Our focus is on the effect of weight changes on the deep learning based mammographic density prediction model pVAS (9). The pVAS method was trained on average expert reader estimates of percent density recorded on Visual Analogue Scales (VAS) (5) and has a strong association with breast cancer risk. For example, a previous study showed that the odds ratio of developing breast cancer between the highest and lowest density quintiles is 4.16 (95% CI 2.53 to 6.82) (9). However, it is not known how pVAS varies with changes in weight. This information may guide how pVAS is used in the future, including whether pVAS should be adjusted based upon weight and weight change.

As a comparison, we investigate how density prediction varies with weight change for a second automated mammographic density method Volpara®Density™ (Volpara Health, Wellington, New Zealand), denoted by Volpara throughout the rest of the paper, which uses imaging physics to compute volumes of dense and fatty tissue. Volpara also has a strong relationship to breast cancer risk with odds ratios between highest and lowest density quintiles of 2.42 (95% CI 1.56-3.78) using the same test set that was used to evaluate pVAS (5).

## 2) Methods and materials

The data for this study are from a recently published weight loss trial (14) which tested three different weight loss interventions amongst 210 women with overweight/obesity enrolled across three UK family history clinics; Manchester University Foundation Trust (MFT), University Hospital Southampton (UHS) and Tameside and Glossop Integrated Care NHS Foundation Trust (T&GICFT) (14). The inclusion criteria for this mammographic density sub-study were that body weight was recorded and raw (for processing) mammograms were available both at baseline and at 12 months. We included pre- and post-menopausal women, but excluded women if they had changes in hormonal factors which could exert weight-independent effects on mammographic density: for example if they became postmenopausal, started endocrine therapy, stopped hormone replacement therapy (HRT) or stopped oral contraceptives during the 12 month period.

Weight at baseline and 12 months was assessed using calibrated scales in the three recruiting centres. Measurements at baseline and 12 months were conducted on the same scales for each trial participant.

The pVAS model was trained using labelled data, with labels generated by expert reader assessment of percent density on visual analogue scales (VAS). The VAS approach to density assessment requires an experienced mammogram reader to assign a percentage density value to each mammographic view by marking with a pen on 10cm VAS scales labelled 0 at one end and 100 at the other. An average of the scores of two independent readers across all mammographic views is taken as the final VAS score (5). This method relies on subjective assessment of the images by experts. Using a matched case-control analysis of mammograms where which had been read as cancer-free, where the cases were subsequently found to have cancer either in a screening interval or at a subsequent screen, the VAS density method outperformed other methods evaluated for risk prediction, and was significantly more predictive than methods assessing purely the quantities of dense and fatty tissue in the breast (5). This improvement may be due to an appreciation by the expert readers of the pattern and location of density in the images in addition to an assessment of the quantity of dense fibroglandular tissue.

The pVAS software (9) uses an artificial intelligence approach with a convolutional neural network trained on a dataset of around 40,000 sets of mammograms from the Predicting Risk Of Cancer At Screening (PROCAS) study (15); it is trained to predict average VAS score for each mammographic view. It produces predictions that give comparable breast cancer risk prediction results to VAS (9). As with VAS assessment, pVAS generates a score per mammographic view, with the final score an average across views and breasts. Whilst pVAS can be used for either raw (‘for processing’) or processed (‘for presentation’) mammograms, in this study raw image data were used. At present pVAS is only available for images from GE HealthCare mammography systems.

Volpara infers the volumes of fibroglandular and fatty tissue from the X-ray attenuation of those tissues coupled with the known physics of the mammography machine and mammographic parameters (8). Volumetric density is assessed as the ratio of the fibroglandular tissue volume to the sum of the fibroglandular and fatty tissue volumes (breast volume). It requires raw (‘for processing’) digital mammographic images and produces an output for each mammographic view. These are averaged to generate a ‘per woman’ density score.

Weight and mammographic images were available at both time points for 109 women in the dietary intervention study. Of these, 43 were excluded from this sub-study as they had confounding hormonal factors, leaving 66 datasets available for analysis. Due to missing data, 61 sets of density estimates for Volpara were available. A further 15 datasets were excluded from analysis by pVAS as the images were not taken on GE HealthCare mammographic systems at both time points. This resulted in 46 sets of images with both pVAS and Volpara density outputs,

We report Spearman rank correlation coefficients between the MD related values and reduction in weight for all participants. For the Volpara scores we consider the volumes of fibroglandular tissue, fatty tissue and volumetric breast density. For the reductions in weight we use both absolute changes (‘Reduction in Weight’) and percentage reduction in weight (weight at first time point minus weight at second time point divided by weight at first time point expressed as a percentage). Uncertainties are all shown at the 95% level.

We report a cross tabulation of those decreasing or increasing weight with decreased or increased mammographic density in Table. We also show plots demonstrating the changes in mammographic density and weight for individual participants for pVAS (Figure 1) and Volpara (Figure 2).

**Figure 1.**
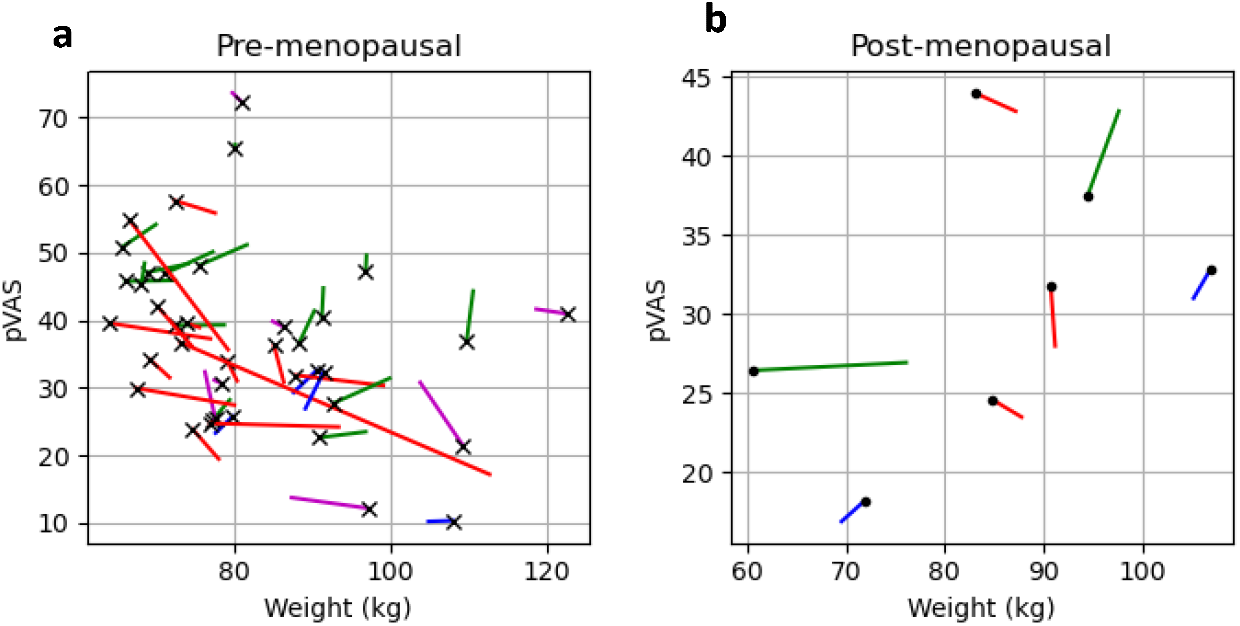
Plots showing variation in pVAS and change in absolute weight for a. pre-menopausal (n=39) b. post-menopausal (n=7) women. The cross (premenopausal) or dot (post-menopausal) represents the second time point and the start of the line the first time point. The different colours represent the change in pVAS and weight blue represents an increase in both pVAS and weight (line towards top right); red an increase in pVAS and reduction in weight (line towards top left); magenta an increase in weight and reduction in pVAS (line towards bottom right); green a reduction in pVAS and weight (line towards bottom left).

**Figure 2.**
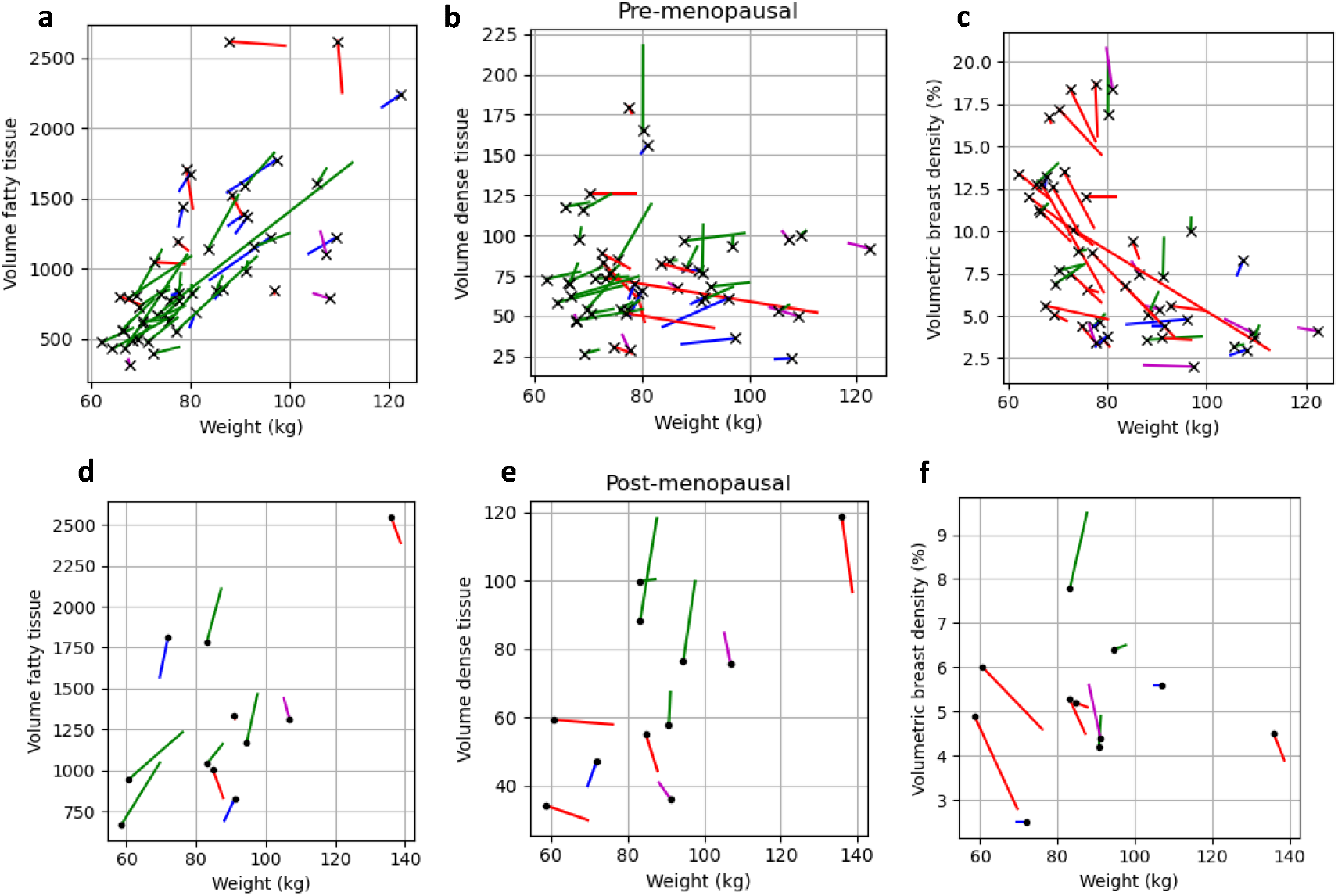
Plots showing variation in the three Volpara measures (volume of fatty tissue, volume of dense tissue, volumetric breast density) and change in absolute weight. Figure a, b, c are for 50 premenopausal women and Figure d, e, f are for the 11 post-menopausal women. The dot (post-menopausal) or cross (premenopausal) represents the second time point and the start of the line the first time point. The different colours represent the change in the measures and weight blue represents an increase in both measure and weight (line towards top right); red an increase in measure and reduction in weight (line towards top left); magenta an increase in weight and reduction in measure (line towards bottom right); green a reduction in the measure and weight (line towards bottom left); Figure a, d Volume of fatty tissue versus weight. Fig b, e Volume of dense tissue versus weight. Fig c, f Volumetric breast density versus weight.

## 3) Results

For the 46 participants included in the pVAS analysis, the mean (standard deviation) age at baseline was 46.1 (4.8), BMI was 32.2 (4.6), and 41 participants were white British, 2 Asian Indian, 1 black.

Caribbean and 1 black other. For the 61 participants included in the Volpara analysis the age at baseline was 46.1 (5.3), BMI was 32.0 (5.2), and 56 participants were white British, 2 Asian Indian, 1 black Caribbean and 1 black other.

Table 1 reports the Spearman rank correlation coefficients for the pVAS scores and the Volpara density measures compared to the absolute reduction in weight (kg) and percentage reduction in weight. Weight reduction does not have a significant correlation with change in pVAS (p=0.27), although the point estimates do show a weak positive correlation (reduction in weight leading to an increase in pVAS). Reduction in weight has no correlation with the Volpara dense tissue volume (p=0.64) while reduction in weight has a strong negative correlation with fatty tissue volume (reduction in weight is associated with a reduction in fatty tissue volume). Therefore reduction in weight has a positive correlation with Volpara volumetric breast density (as weight falls volumetric breast density increases). The uncertainties are large due to the relatively small amount of data.

**Table 1.**
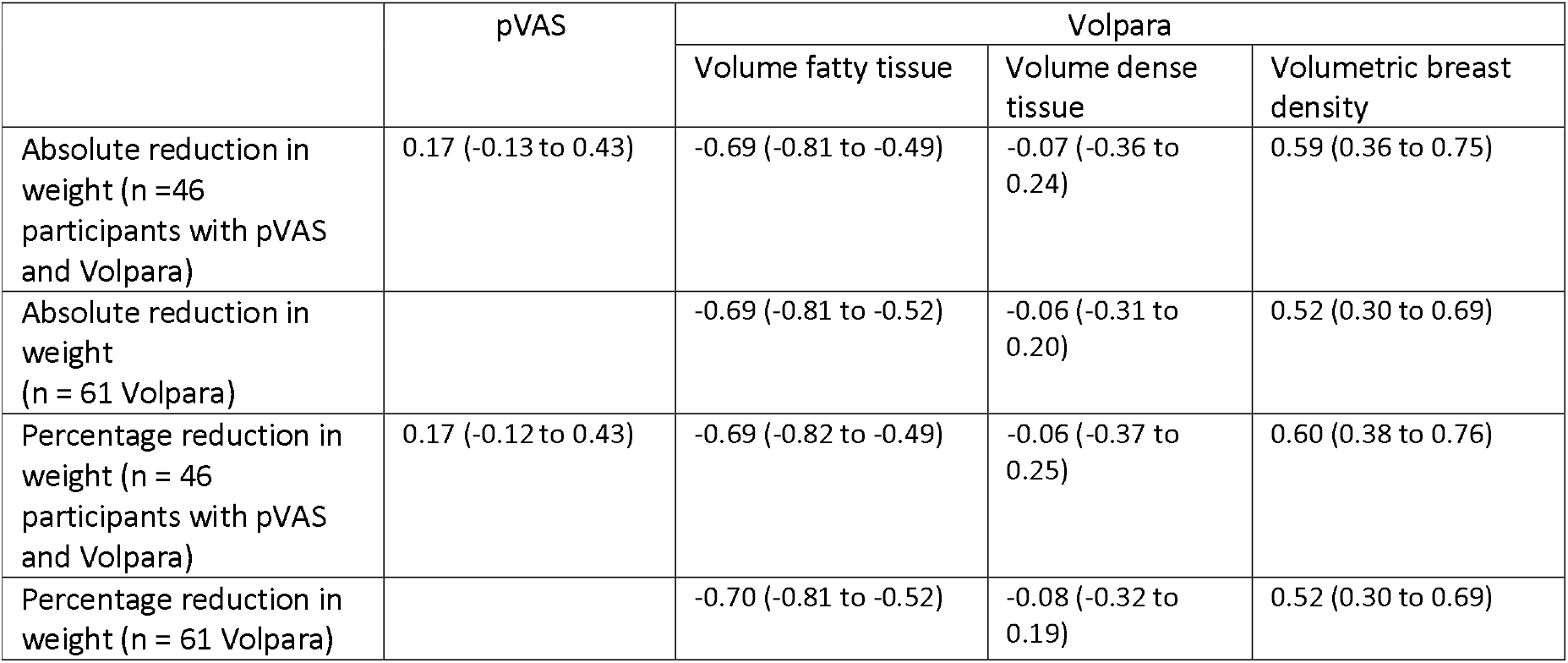
Spearman rank correlation coefficients for pVAS scores and Volpara scores (volume fatty tissue, volume dense tissue and volumetric breast density) compared to reduction in absolute weight and percentage reduction in weight.

|  | pVAS | Volpara |  |  |
| --- | --- | --- | --- | --- |
|  |  | Volume fatty tissue | Volume dense tissue | Volumetric breast density |
| Absolute reduction in weight (n = 46 participants with pVAS and Volpara) | 0.17 (-0.13 to 0.43) | -0.69 (-0.81 to -0.49) | -0.07 (-0.36 to 0.24) | 0.59 (0.36 to 0.75) |
| Absolute reduction in weight (n = 61 Volpara) |  | -0.69 (-0.81 to -0.52) | -0.06 (-0.31 to 0.20) | 0.52 (0.30 to 0.69) |
| Percentage reduction in weight (n = 46 participants with pVAS and Volpara) | 0.17 (-0.12 to 0.43) | -0.69 (-0.82 to -0.49) | -0.06 (-0.37 to 0.25) | 0.60 (0.38 to 0.76) |
| Percentage reduction in weight (n = 61 Volpara) |  | -0.70 (-0.81 to -0.52) | -0.08 (-0.32 to 0.19) | 0.52 (0.30 to 0.69) |

**Table 2.**
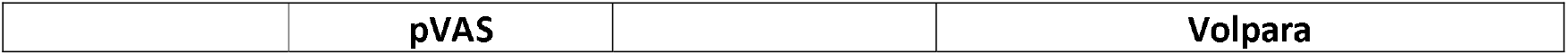
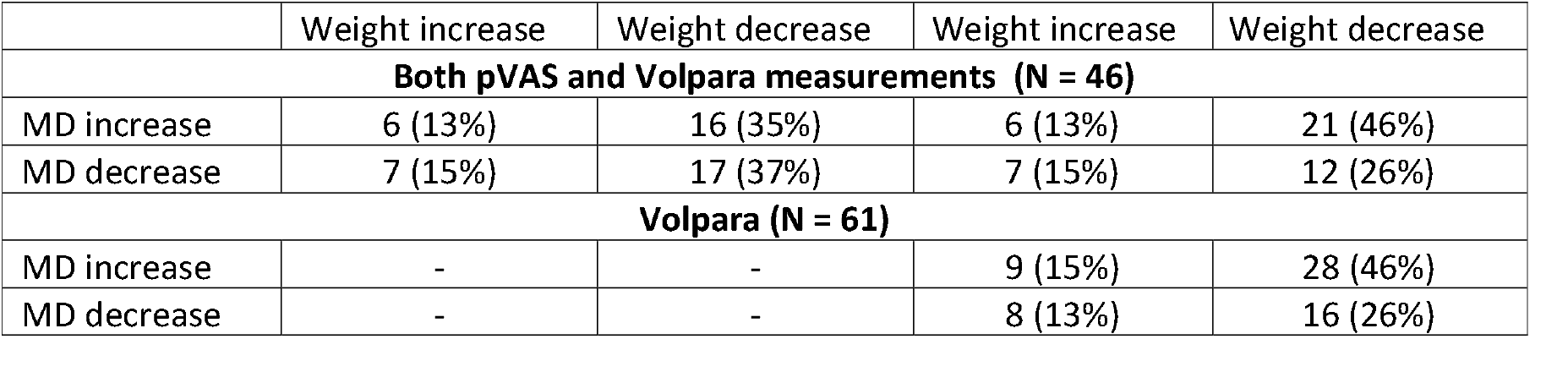
Numbers of participants with weight increase/decrease and increasing/decreasing mammographic density measures. Percentage in brackets.

|  | Weight increase | Weight decrease | Weight increase | Weight decrease |
| --- | --- | --- | --- | --- |
| **Both pVAS and Volpara measurements (N = 46)** |  |  |  |  |
| MD increase | 6 (13%) | 16 (35%) | 6 (13%) | 21 (46%) |
| MD decrease | 7 (15%) | 17 (37%) | 7 (15%) | 12 (26%) |
| **Volpara (N = 61)** |  |  |  |  |
| MD increase | - | - | 9 (15%) | 28 (46%) |
| MD decrease | - | - | 8 (13%) | 16 (26%) |

Table shows the number of participants who had a weight increase or decrease and MD increase or decrease for those with both pVAS and Volpara (n=46) and Volpara (n=61). Of the 46 participants with both measures 33 had a decrease in weight and 13 an increase. For Volpara, when there was a decrease in weight, more women had an increase in mammographic density, while for pVAS, when there was a decrease in weight, half of the participants had an increase in MD and half a decrease in MD.

In Figure 1 we show plots of the change in pVAS and absolute weight between the baseline and 12 months for 39 premenopausal women(Figure 1a) and 7 post-menopausal women (Figure 1b). The mean (standard deviation) of the baseline and 12 months weights are 86.0 kg (12.2 kg) and 82.5 kg (13.8 kg) respectively. The mean (standard deviation) of the pVAS scores at baseline and 12 months are 35.8% (13.0%) and 36.3% (12.4%) respectively.

In Figure 2 we show plots of the three Volpara measures (fatty, dense and % density) against absolute weight in kilograms. Figure 2a, b, c are the 50 premenopausal women, Figure 2d, e, f are the 11 post-menopausal women. The mean (standard deviation) of the baseline and 12 months weights are 86.0 kg (14.0 kg) and 82.7 kg (15.6 kg) respectively. The mean (standard deviation) of the Volpara density scores at baseline and 12 months are 7.3% (4.3%) and 7.9% (4.5%) respectively.

## Discussion

We have demonstrated that when density is measured using pVAS, there is no statistically significant correlation between weight change and density change. In contrast our analysis reconfirms previous reports of the inverse correlation between change in weight and volumetric breast density as measured by Volpara which is driven by a reduction in the volume of fatty tissue in the breast with weight loss.

The pVAS model was trained on VAS from expert reader assessment which are subjective measures of MD. The pVAS density measure does not appear to be strongly dependent on the ratios of tissue types, and it is possible that other aspects of breast density such as pattern and location contribute to the expert assessments. These may be less likely to be affected by weight change. This is the first study to investigate the effect of weight change on pVAS and it appears that as pVAS is less affected by changes in weight that there may be no need to adjust for BMI.

A strength of this study is the contemporaneous measures of weight, pVAS and Volpara. Limitations include the relatively small sample size which results in large uncertainty intervals; which limits the conclusions we can draw, particularly with regard to whether changes in dense tissue volume or pVAS are related to weight change.

## Conclusions

pVAS percent density, trained on VAS scores, shows no statistically significant correlation with change in weight. The lack of association between weight change and change in pVAS suggests it is not a useful biomarker of the potential reduction in risk of breast cancer alongside weight loss. Also that measurements of pVAS for predicting risk may not need adjustment for BMI, although this would need to be confirmed in a larger dataset.

## Data Availability

All datasets used and analysed during the current study are available on reasonable request.

## Acknowledgements

The project was funded by Prevent Breast Cancer (registered charity number 1109839) and supported by the NIHR Manchester Biomedical Research Centre (IS-BRC-1215-20007) infrastructure. The funders had no role in the design, conduct, analysis or write up of the study. MH, DGE, AH, SA are supported by the NIHR Manchester Biomedical Research Centre (IS-BRC-1215-20007).

The views expressed are those of the authors and not necessarily those of the National Health Service, the National Institute for Health Research, or the Department of Health and Social Care.

